# Early marriage, preterm birth and school dropout: an intergenerational cycle of risk?

**DOI:** 10.1101/2025.07.23.25332058

**Authors:** Jonathan C Wells, Qisty Noviyanti, Akanksha A Marphatia, Emeline Rougeaux

**Author notes:** Corresponding author: Jonathan Wells [1].

## Abstract

**Background:** Across generations, girls’ early marriage recurs in high-risk groups, however there is poor understanding of how behaviour and biology interact in this context. We hypothesised an intergenerational cycle of risk, linking early marriage, preterm birth and school dropout, and evaluated the evidence in low-/middle-income countries.

**Methods:** We conducted a systematized review, searching articles published from 2000-2025. We tested the hypotheses: H1 that early marriage is associated with preterm birth; H2 that preterm birth is associated with low educational attainment; and H3 that school dropout is associated with early marriage. Hypothesis-specific search terms and eligibility criteria were applied. We also tested the hypothesis (H4) that preterm birth is associated with reduced cognitive function, evaluating systematic reviews of research from any setting. We identified a total of 185 empirical articles for H1-3, with 26 satisfying the criteria for full review, and 5 systematic reviews for H4.

**Results:** The available empirical evidence consistently supported H1-3, though there were fewer studies for H1 (4 studies) and H2 (2 studies) compared to H3 (20 studies). The systematic reviews demonstrated strong evidence for H4.

**Conclusions:** Our reviews support the hypothesis of an intergenerational cycle of risk linking early marriage, preterm delivery and low educational attainment. At a societal level, the cycle is perpetuated when adolescent girls leave school and marry early. Mechanistically, early marriage may increase risk of preterm birth through psychosocial stress and early childbearing, while preterm birth undermines schooling through cognitive impairment. Interventions to prevent early marriage may help disrupt the cycle.

## Introduction

Early childbearing is detrimental to maternal and child health (Fall et al., 2015; Marphatia, Ambale, & Reid, 2017), but is positively correlated with fertility (Raj, Saggurti, Balaiah, & Silverman, 2009; Sagalova, Nanama, Zagre, & Vollmer, 2021). From an evolutionary perspective, this indicates a trade-off between life history traits, whereby increased allocation of resources to reproduction in women comes at the expense in their investment in growth and maintenance (Marphatia, Saville, Manandhar, Cortina-Borja, Reid, et al., 2021). This trade-off may also be subject to sexual conflict over the timing of the onset of reproduction, as fathers gain the fitness benefits of women reproducing early, without themselves paying the health costs (J.C. Wells, 2022).

In many countries, particularly in South Asia, early reproduction is preceded by high rates of early marriage, as reproduction outside marriage is socially unacceptable (Marphatia et al., 2017). Early marriage recurs across generations in high-risk groups (Marphatia, Wells, Reid, Bhalerao, & Yajnik, 2024), and may be part of a broader inter-generational cycle of disadvantage that embraces poverty, malnutrition, gender inequality, early reproduction and risky behaviour (J.C. Wells et al., 2019). Early marriage has detrimental effects on women beyond early reproduction, and is associated with a range of adverse outcomes including lower schooling, low autonomy and intimate partner violence (Kidman, 2017; Marphatia et al., 2017; Subramanee et al., 2022). It is therefore the target of extensive policy efforts (Malhotra & Elnakib, 2021b; UNICEF, 2023).

More broadly, intergenerational associations have been documented for a range of adverse socio-economic and human capital outcomes. For example, low parental education predicts poorer schooling outcomes in the offspring (Marphatia et al., 2016; Marphatia, Reid, & Yajnik, 2019), while poverty also tracks across generations (Harper, Marcus, & Moore, 2003; Van Ryzin, Fishbein, & Biglan, 2018). In the 1960s, Oscar Lewis presented a ‘culture of poverty’ hypothesis, proposing that the perpetuation of adverse socio-economic outcomes across generations was partly driven by the transmission of cultural values (Lewis, 1966). In the US, this theory stimulated a ‘war on poverty’ with the aim of ‘correcting’ such values. When the programmes, which made little effect to change structural factors, did not succeed, the intractability of the ‘culture of poverty’ was invoked as the explanation – its intergenerational ‘cultural basis’ had seemingly made poverty ineradicable (Seligman, 1968).

Although Lewis encouraged policy makers to view behaviours as learned and culturally transmitted, they might also be sensitive to physiological mechanisms and biological exposures. Among the traits that Lewis considered characteristic of the culture of poverty are several that are highlighted in evolutionary life history theory, including low life expectancy, early initiation into sex and high mortality risk (Lewis, 1966; Promislow & Harvey, 1990). Over recent decades, the notion of behaviour and physiology interacting dynamically across intergenerational timescales has attracted increasing attention. For example, there is growing interest in how psychosocial stressors and material inequalities ‘get under the skin’ and impair health and development across generations (Cheng, Johnson, & Goodman, 2016; Entringer et al., 2011; Yehuda et al., 2016). Exposure to pollutants and toxins may exert similar effects (Lowell, Morie, Potenza, Crowley, & Mayes, 2022; Schell, 1997).

For early marriage, the primary driving factor is widely assumed to be household poverty (Psaki et al., 2021), and this has led to cash-transfer programmes aiming to delay girls’ marriage. These programmes have generally had low efficacy (Malhotra & Elnakib, 2021a). However, a major limitation of many studies that linked early marriage with poverty is that economic assets were measured only in households where the woman was already married, thus indexing marital rather than natal household wealth. In a rare study where assets were measured in the natal household, girls’ low education was a robust predictor of age at marriage, whereas independent of that, household assets showed no association (Marphatia, Saville, Manandhar, Cortina-Borja, Wells, et al., 2021).

In South Asian societies, marriage decisions are also strongly influenced by socio-cultural norms relating to a range of factors, including cementing family ties, dowry payments and ensuring brides’ chastity and subservience (Fattah & Camellia, 2022; UNFPA, 2019). In India, socio-cultural norms for marriage age are changing more slowly than those for education (Marphatia, Wells, Reid, Poullas, et al., 2024). However, if marriage decisions are considered entirely the product of such norms, then difficulties in changing marriage practices might appear to be reiterating the ‘culture of poverty’ argument - that early marriage is a cultural practice that is simply ‘too engrained’ to change.

Here, we draw on emerging evidence linking early women’s marriage with the risk of adverse physical outcomes in the offspring, to generate a new overarching hypothesis: that the perpetuation of early marriage across generations involves the interaction of behavioural decisions with physiological traits, contributing to a complex multi-mechanism intergenerational cycle of risk. By specifying the consecutive risks, we might be able to reduce the intergenerational propagation of disadvantage. Our recent research in lowland rural Nepal found that independent of age at first childbearing, girls’ early marriage was associated with an increased risk of delivering a preterm infant (Miller et al., 2022). In the same cohort, we had also shown that girls’ school dropout was associated with their early marriage (Marphatia, Saville, et al., 2019). Given that in a high-income setting, shorter gestation length showed a dose-response association with difficulties in school (MacKay, Smith, Dobbie, & Pell, 2010), it seemed possible that early marriage, preterm birth and poor schooling might reinforce each other across generations, and hence form one component of the intergenerational cycle of disadvantage. From an evolutionary life-history perspective, exposures that undermine maternal investment in daughters’ longevity may favour an earlier shift of daughters’ metabolic resources to reproduction (Griskevicius, Tybur, Delton, & Robertson, 2011; J.C. Wells et al., 2019). In contemporary human settings, such interactions may be mediated by both physiological mechanisms and cultural institutions such as school dropout and early marriage.

This component of the cycle of disadvantage is therefore projected to incorporate three specific risks, framed here as hypotheses (H) and illustrated in **Figure 1**:

H1. Girls’ early marriage is associated with increased risk of delivering a preterm infant
H2. Preterm birth compromises educational attainment of the offspring, increasing the risk of school dropout
H3. Poor educational attainment and school dropout increase the likelihood of girls’ early marriage

**Figure 1.**
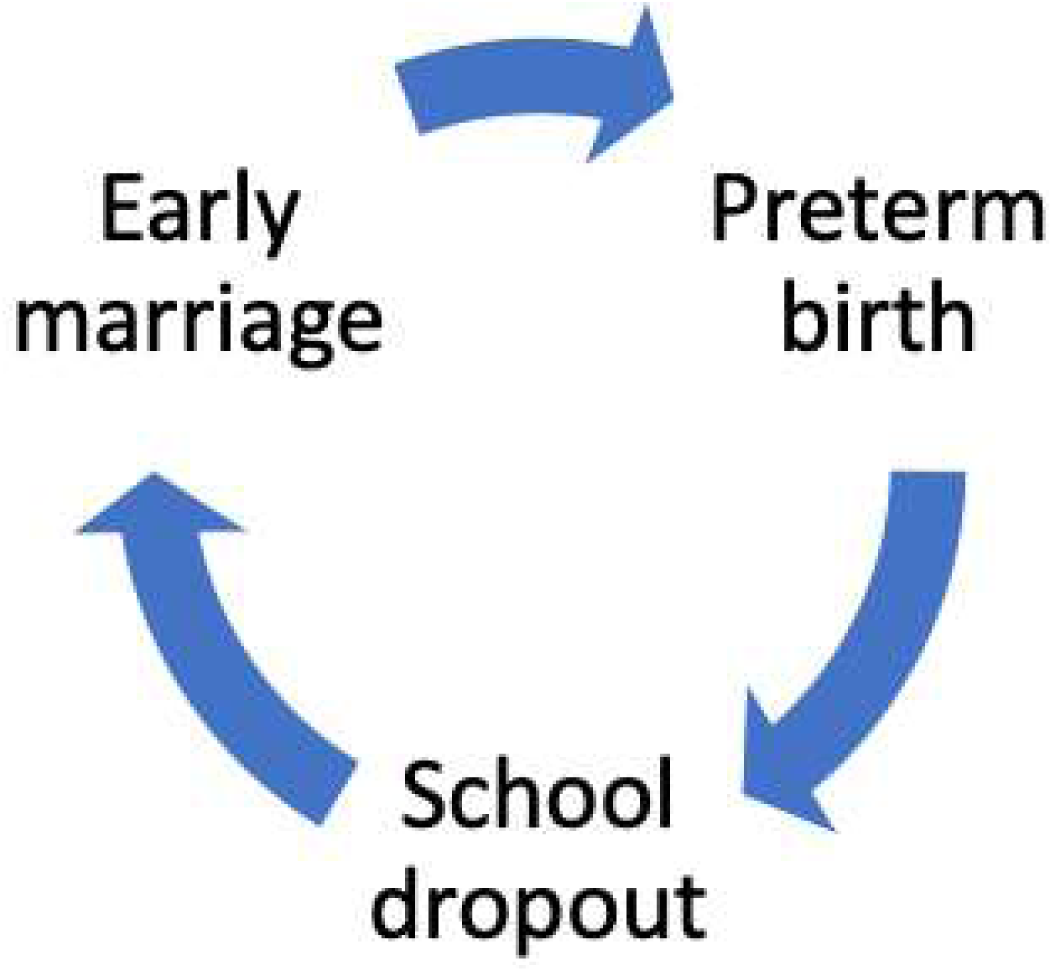
Schematic diagram of the hypothesised intergenerational cycle, linking girls’ early marriage, preterm birth and school dropout. The associations are not deterministic at the individual level, rather at the population level, each outcome in the cycle increases the probability of the next outcome occurring.

Whether epidemiological evidence supports these hypotheses in low- and middle-income countries (LMICs), where early marriage is especially common (Marphatia et al., 2017), remains unclear. We therefore conducted systematic literature reviews to test these hypotheses. As prospective longitudinal studies of cognitive function in school children in LMICs remain sparse, we also searched for systematic reviews of research from any setting, to test a further hypothesis relating to underlying mechanisms:

H4: Preterm birth is associated with reduced cognitive function in school-aged children.

## Methods

Given the complexity of our overarching hypothesis, we conducted a systematized review (Grant & Booth, 2009), in order to link together the results of several systematic searches with specific eligibility criteria into a broader synthesis.

### Search strategy

The literature searches were conducted using the electronic databases Scopus, PubMed, and Google Scholar in March 2025, considering studies published in 2000 to 2025 in English. For each hypothesis, broad search terms were applied to both titles and abstracts to increase the likelihood of identifying relevant articles. The search terms were as follows:

*H1: Early Marriage and Preterm Birth*: (“Early marriage” or “child marriage” or “married girls” or “adolescent marriage” or “child and adolescent marriage” or “married adolescent” or “underage marriage” (and not “adolescent pregnancy” or “early childbearing”)) and (“preterm birth” or “preterm delivery” or “premature birth”).

*H2: Preterm Birth and Educational Attainment*: (“Preterm birth” or “preterm delivery” or “premature birth”) and (“schooling” or “school dropout” or “education” or “educational attainment” or “educational status” or “educational level” or “educational achievement” or “educational outcome” or “educational disadvantage” or “age at school entry” or “school grade” or “school level” or “school performance” or “schooling attainment”)

*H3: Educational Attainment and Early Marriage*: (“schooling” or “school dropout” or “education” or “educational attainment” or “educational status” or “educational level” or “educational achievement” or “educational outcome” or “educational disadvantage” or “age at school entry” or “school grade” or “school level” or “school performance” or “schooling attainment”) and (“early marriage” or “child marriage” or “married girls” or “adolescent marriage” or “child and adolescent marriage” or “married adolescent” or “underage marriage” and not (“adolescent pregnancy” or “early childbearing”)).

*H4:* H4 included the same search terms as H2 but searched for systematic reviews of studies in any setting. Broader school and education search terms were retained as research on school performance and educational attainment frequently includes approaches and scales reflecting cognitive skills and development (such as literacy and intelligence quotient). The inclusion criteria differed from H2 however, as detailed in the subsequent section.

### Inclusion and Exclusion Criteria

For H1-3, only empirical studies conducted in LMICs with a quantitative design study were eligible. If mixed-method studies were identified, only quantitative findings were analysed. LMICs were categorised using the 2022 World Bank definition of countries with gross national income under $13,205 (World Bank, 2022). All countries within the low, lower-middle, and upper-middle group fell into this category.

Articles were only included if they investigated the association between early marriage and preterm birth (H1), preterm birth and educational attainment/school performance/school dropout (H2), and educational attainment/school performance/school dropout and early marriage (H3). Educational attainment was defined as the number of years completed in school, while school dropout was defined as leaving school prior to completion of specified curriculum requirements. No strict definition was applied to define school performance, as a wide range of assessments exists worldwide. Early marriage was defined as a formal or informal union between a girl-child under 18 years old, with no restriction on age of the spouse (UNICEF, 2018).

Preterm birth was defined as a live birth below 37 weeks of gestation (Blencowe et al., 2012). For H1 and H2, given limited evidence, no restriction was applied to degree of preterm delivery (extremely, very, and late preterm). However, the majority of evidence for both hypotheses relates to later preterm deliveries, hence our review reflects this. First, the burden of late preterm infants is significantly higher in LMICs compared to HICs (March of Dimes, PMNCH, Save the Children, & WHO, 2012). Second, late preterm infants have higher survival rates compared to very or extremely preterm infants in LMICs, due to limited availability of medical care (>50% vs 10%) (Blencowe et al., 2013). This is relevant to our interest in life-course outcomes for the other two hypotheses. Third, very or moderate preterm children appear to have distinct educational outcomes compared to late preterm children (Loftin et al., 2010; Smyrni et al., 2021).

For H4, only systematic reviews of quantitative studies were included, with no restriction on country in the search operation. Within these we then specifically assessed whether studies explored associations of preterm birth with cognitive outcomes.

Articles were excluded if they: had a qualitative design, were grey literature, discussed genetic traits as predisposing factors, focussed on early marriage in boys, and/or focussed on early childbearing without marriage.

### Data Extraction

Once a study was considered to satisfy eligibility criteria, it was subjected to analysis and the findings were summarised. For H1-H3, data were extracted for the following variables: sample size, demographic characteristics, country, operational definition for exposure and outcome, adjusted factors, and main findings. For H4, data were extracted for the following variables: number of studies, setting, operational definition for exposure and outcome, risk of bias assessment, and main findings.

## Results

Across hypotheses 1 to 3, a total of 185 empirical articles were found and 26 articles satisfied the criteria for full review.

### Study Characteristics

For H1, there were four studies from Nepal, India, Brazil and Ecuador. The studies from Brazil and Ecuador were national studies covering all live births to women below 25 years in 2011-2018 (Brazil) and 2014-2018 (Ecuador) (M. L. Urquia et al., 2022; Urquia, Batista, Cunha Cardoso, Grandi, & Fafard St Germain, 2022). One of these studies also included data from the USA and Canada but these are not reported here. The study from India was a small study of 158 married couples in a village in Gujarat, India (Pandya & Bhanderi, 2015). The study from Nepal assessed a sample of close to 18,000 married girls/women aged 10-49 years from a trial conducted in rural lowland Nepal (Miller et al., 2022). These studies all explored associations of early marriage (either defined as marriage below 18 years, or at specific ages below 18, or using marital status alongside maternal age) with preterm birth. One study did not define preterm birth by gestational age (Pandya & Bhanderi, 2015), and one study distinguished sub-types of preterm status (Marcelo L. Urquia et al., 2022).

For H2, two studies were identified. The first assessed associations of preterm birth with educational attainment in 4,518 adults across five countries (Brazil, Guatemala, India, the Philippines, South Africa) (Stein et al., 2013). The second assessed associations of gestational age (but not preterm birth) and secondary school dropout in 700 children in India (Marphatia, Reid, et al., 2019).

For H3, twenty studies were identified analysing data from 31 countries, spanning sub-Saharan Africa (25 countries) (Bengesai, Amusa, & Makonye, 2021; Bhan et al., 2019; Fang et al., 2024; Glick, Handy, & Sahn, 2015; Glynn et al., 2018; Lami et al., 2023; Sagalova et al., 2021; Zegeye et al., 2021), Asia (5 countries) (Bhan et al., 2019; Cameron, Contreras Suarez, & Wieczkiewicz, 2023; Kanji, Carmichael, Darko, Egyei, & Vasilakos, 2024; Kumar, Patel, Debbarma, & Saggurti, 2023; Y. Liang & Yu, 2022; Marphatia, Reid, et al., 2019; Paul, 2019, 2020; Prakash et al., 2017; Roy et al., 2021; Sekine & Hodgkin, 2017; Singh, Shekhar, & Gupta, 2024), and South America (1 country) (Bhan et al., 2019). Not every study was analysed independently, for example one study pooled data from 21 sub-Saharan African countries (Sagalova et al., 2021), while another study pooled data from India, Ethiopia, Vietnam and Peru (Bhan et al., 2019). The most commonly studied country was India (nine studies). Fifteen studies investigated only educational attainment (Bengesai et al., 2021; Cameron et al., 2023; Fang et al., 2024; Glick et al., 2015; Kanji et al., 2024; Lami et al., 2023; Y. Liang & Yu, 2022; Marphatia, Reid, et al., 2019; Marphatia, Saville, et al., 2019; Paul, 2019, 2020; Roy et al., 2021; Sagalova et al., 2021; Singh et al., 2024; Zegeye et al., 2021), three studies discussed mainly school dropout (Bhan et al., 2019; Kumar et al., 2023; Prakash et al., 2017; Sekine & Hodgkin, 2017), and one study investigated both educational attainment and school performance (using age for grade) and school drop-out (Glynn et al., 2018).

For H1 to H3, the sample size varied from 158 to 7,953,376, since many studies used national demographic or health survey data. For H4, five systematic reviews were identified, summarising the results of a total of 217 original studies.

## Main findings

### H1. Girls’ early marriage is associated with increased risk of delivering a preterm infant

**Table 1** summarises the study characteristics and main findings for the studies on early marriage and preterm birth (H1) (Miller et al., 2022; Pandya & Bhanderi, 2015; M. L. Urquia et al., 2022; Marcelo L. Urquia et al., 2022). All four studies reported an association of early reproduction with an increased risk of having a preterm delivery, though the magnitude of effect varied by setting, and one study found that the effect was only significant in primigravidae. The two South American studies focused on age at reproduction as the primary exposure, however the reported data allowed calculation of the associations for early marriage. In both Ecuador and Brazil, earlier marriage was associated with increased odds of preterm birth, with the earlier the marriage, the greater the risk. In the two South Asian studies, where marriage is a near-universal precedent of childbearing, earlier age at marriage was again associated with increased risk of preterm delivery in dose-response manner. In the study from Nepal, the association was only apparent in first-time mothers, but was independent of the age at childbearing.

**Table 1.** Summary of studies on early marriage and preterm birth.

| Author (year) | Country | Sample | Exposure/outcome | Adjusted factors | Findings |
| --- | --- | --- | --- | --- | --- |
| Miller et al. (2021) | Nepal | 17,974 married and conceived women aged 10-49 years | PB: <37 weeks/ 259 days<br>EM: < 14 years, 15 years, and 16-17 years. | Maternal education, caste group, material assets, maternal nutritional status (BMI, MUAC, Height). | Primigravidae married <14 years greater likelihood of PB (OR: 1.28 [CI: 1.01, 1.62]); no significant result for girls married at 15 years (1.12 [0.89, 1.41]) or 16-17 years (1.12 [0.90, 1.39]). No significant association in multigravidae. |
| Pandya & Bhandari (2015) | India | 158 couples | PB: Not defined. Based on records such as Mamta Card or self-response<br>EM: < 18 years from self-report/marriage certificate | None. | Prevalence of PB higher in women married <18 years compared to married 18+ years (OR: 4.74 [CI:1.01, 22.36]*). No further analyses carried out. |
| Urquia et al. (2022) | Brazil | 7,953,739 births to mothers <25 years | PB: very preterm (24–31 weeks) and moderately preterm (32–36 weeks)<br>EM: 16–17 years and ≤15 years. | Father's age, late prenatal care initiation, maternal education, year of birth, infant sex | Compared to married 20-24 year old women: women/girls married at 18-19 years, 16-17 years and <16 years had higher odds of very PB (OR [CI]: 1.09 [1.04, 1.15], 1.44 [1.33, 1.56], 2.13 [1.71, 2.67])*; and women/girls married at 18-19 years, 16-17 years and <16 years had higher odds of moderate PB (OR [CI]: 1.12 [1.10, 1.14], 1.39 [1.36, 1.43], 1.76 [1.63, 1.91])* |
| Urquia et al. (2022) | Brazil, Ecuador, (USA, Canada but not included here) | Brazil: 24,446,170 births, Ecuador: 1,581,550 births to mothers <25 years | PB: <37 weeks gestation<br>EM: assessed from marital status and maternal age at the time of birth. | Infant sex, previous birth, year of birth, maternal ethnicity.<br>Brazil only: paternal age, prenatal care initiated in 1<sup>st</sup> trimester, state, age-appropriate low education.<br>Ecuador only: foreign-born mother, adequacy of the number of prenatal care visits for gestational age (WHO), maternal literacy, maternal region of residence and rurality. | In Brazil, compared to married 20-24 year old women, women/girls married at 18-19 years and <18 years had higher odds of PB (OR [CI]: 1.12 [1.10, 1.14] and 1.43 [1.40, 1.47] respectively)*.<br>In Ecuador, compared to married 20-24 year old women, women/girls married at 18-19 years and <18 years had higher odds of PB (OR [CI]: 1.08 [1.00, 1.17] and 1.13 [0.98, 1.30] respectively)*. |

Abbreviations: EM: Early Marriage, PB: Preterm Birth, BMI: Body Mass Index, MUAC: Mid-Upper Arm Circumference, OR: Odds Ratio, CI: 95% Confidence Interval.
\*Crude OR and 95% CI estimated from crude numbers reported in the study

### H2. Preterm birth compromises educational attainment of the offspring

**Table 2** summarises the study characteristics and main findings for the association of preterm birth and educational attainment/ academic performance/ school dropouts (H2) (Marphatia, Wells, Reid, & Yajnik, 2022; Stein et al., 2013). A study of 700 adolescents from India found an inverse association of gestational age with the likelihood of school dropout. Each additional week of gestational age was associated with 13% lower odds (95% CI 1, 23) of dropout. A pooled analysis of five birth cohorts from Guatemala, Brazil, South Africa, India and the Philippines found that compared with adults born term, those born preterm had 0.44 years lower (95% CI, 0.17-0.71 years) educational attainment.

**Table 2.** Summary of studies on preterm birth and educational outcomes.

| **Author (year)** | **Country** | **Sample** | **Exposure/outcome** | **Adjusted factors** | **Findings** |
| --- | --- | --- | --- | --- | --- |
| Marphatia et al. (2022) | India | 700 children | Gestational age (duration of pregnancy) (PB not assessed)<br>School dropout during secondary school | Child age, sex, growth and early education and maternal and paternal education, household socio-economic status. | Greater gestational age was associated with a 13% lower odds of secondary school dropout (OR [CI]: 0.87 [0.77, 0.99]). |
| Stein et al. (2013) | Brazil, Guatemala, India, the Philippines, and South Africa. | 4,518 adults | PB: <37 completed weeks of gestation<br>Educational attainment: completed years of schooling | Site, sex, and age at adult assessment. | Compared with adults born term, educational attainment was 0.44 years lower (CI, 0.17, 0.71 years) in those with PB. |

*Abbreviations: PB: Preterm Birth, OR: Odds ratio, CI: 95% Confidence Interval.*

### H3. Poor educational attainment increases the likelihood of girls’ early marriage

**Table 3** summarises the study characteristics and main findings for the studies on educational attainment, academic performance or school dropout and early marriage (Bengesai et al., 2021; Bhan et al., 2019; Cameron et al., 2023; Fang et al., 2024; Glick et al., 2015; Glynn et al., 2018; Kanji et al., 2024; Kumar et al., 2023; Lami et al., 2023; Y. Liang & Yu, 2022; Marphatia, Reid, et al., 2019; Marphatia et al., 2020; Paul, 2019, 2020; Prakash et al., 2017; Roy et al., 2021; Sagalova et al., 2021; Sekine & Hodgkin, 2017; Singh et al., 2024; Zegeye et al., 2021). All 20studies reported an association of lower education and early marriage, through the nature of the association varied across studies and the direction of the association was analysed in different ways. All fifteen studies that assessed this found that educational attainment was lower among early-married girls, or that increased education was associated with reduced odds of early marriage. All four studies that assessed this found that early-married girls were more likely to have dropped out of school, or that school dropout was associated with increased odds of early marriage. The available evidence indicates that the association of early marriage poor education may be a two-way street, with each a risk factor for the other.

**Table 3.** Summary of studies on educational outcomes and early marriage.

| Author (year) | Country | Sample | Exposure/outcome | Adjusted factors | Findings |
| --- | --- | --- | --- | --- | --- |
| Bengesai et al. (2021) | Zimbabwe | 2,380 ever-married women aged 20-29 years | EM: Age at marriage <18 years<br>Lower secondary school completion (11 years): Yes, no | Place of residence, religion, spouse education (years), type of marital union (monogamy/polygamy/missing), age at sexual debut (including informal union), sex of HH head, spousal age difference, age of respondent | Prevalence of lower secondary school completion 62% lower in EM women (PR [CI]: 0.381[0.30, 0.49]) compared to those married later (≥18 years) |
| Bhan et al. (2019) | India, Ethiopia, Vietnam, Peru | 1338 girls | EM: Age at marriage < 18 years<br>School dropout: dropped out by age 12/15 years vs. stayed in school | Country, wealth quartiles, rural/urban residence, maternal education (none, primary, secondary or higher), whether parents are alive, early menarche, parental beliefs on value of education. | Girls married before 16 years, at 16-17 years and 18-19 years had a greater risk of school dropout by age 12/15 years compared to girls not married by 19 years (RRR [CI]: 11.76 [6.97, 19.8], 4.91 [3.07, 7.86], 4.86 [2.79, 8.46] respectively). Note that no girls married at ages 16-17 and 18-19 years in Ethiopia and it is therefore not included in those results. |
| Cameron, Suarez & Wieczkiewicz (2023) | Indonesia | 40,800 men and women [only women reported here] | EM: Age at marriage < 18 years<br>Educational attainment: total years completed by the time of the survey | #A Urban area, muslim, age, parental education, parent marital status (at age 12 years), and village fixed effects (accounting for differences between individuals in the same villages)<br>#B Urban area, age, and sister fixed effects (controls for all family characteristics, both observable and unobservable, experienced by sisters e.g., family background). | Women who marry before the age of 18 obtained 1.65 years (SD: 0.09) less education after adjusting for #A and 0.91 years (SD: 0.22) less education after adjusting for #B |
| Fang et al. (2024) | Nigeria | Women aged 15-29 years in 42,000 households | EM: Age at first marriage < 18 years<br>Educational attainment: completed secondary education or attained a higher level of education | Women's age in 5-year groups, de facto region of residence, area of residence (urban, rural), current marital status of the respondent, and family poverty (living in households in the lowest wealth quintile). | The marginal effect of child marriage on whether the respondent completed secondary or higher education was -22.98 %. |

**Table 3. Continued**
| Author (year) | Country | Sample | Exposure/outcome | Adjusted factors | Findings |
| --- | --- | --- | --- | --- | --- |
| Glick et al. (2015) | Madagascar | 2,336 females aged 12-25 years | EM: Age at marriage<br>Educational attainment: number of years completed | Year of birth, family background (parent education, parent mortality, assets, religion, HH head ethnicity), and community variables (province indicators, urban residence, electricity, water source, distance to public services, transport, and infrastructure) | Each additional year of education associated with 1.5 year increase median marriage age |
| Glynn et al. (2018) | Malawi | 8,576 girls aged over 12 | EM: Age at first marriage<br>Educational attainment: years overage for grade<br>School drop-out: in primary school or beyond | Parental education, vital status of parents, living arrangements (HH size, number of children under 5 in HH, living with parents), sex of head HH, SES, year of interview | Compared to girls in primary school, rates of marriage across ages 13 to 18 years were cumulatively higher for girls who had dropped out in primary school (eg marriage by 13 years HR 1.79 [CI 1.20, 2.66], marriage by 16 years HR 4.67 [CI 2.98, 7.32]. Similar results for those who dropped out of school beyond primary, eg marriage by 15 years HR 2.30 [CI 1.07, 4.92], marriage by 18 years HR 3.58 [CI 2.17, 5.90]. |
| Kanji et al. (2024) | India | 1,008 children aged 8 up to 22 years | EM: Age at marriage < 18 years<br>Educational attainment: school grade achieved at different ages | None | Lower mean educational attainment found at age 12 (but not 8 years) for girls married before 18 years compared to girls not. Difference in difference analyses show EM negatively affects women's self-reported health and educational attainment by age 22 years (from baseline 8 years). |
| Kumar et al. (2023) | India | 6,178 adolescent girls enrolled in school at survey wave 1 | EM: Married at age 15-19 years<br>School drop-out: dropped out of school between survey waves 1 and 2 | None | Prevalence of school dropout higher in married than unmarried girls at 84.3% vs 45.8% (no further statistical analyses carried out). |
| Lami et al. (2024) | Ethiopia | 990 women of reproductive age | EM: Age at marriage < 18 years<br>Education level: level completed at the time of the survey | Age, religion, husband's educational level, occupation of women, husband's occupation, residence, means of engagement/marriage, consent at marriage, knowing the age of legal marriage, and decision-maker at marriage. | Women who had a diploma and above educational level were 74% [AOR: 0.26 (0.10, 0.70)] less likely to have had a child marriage than those who had a primary school educational level. No associations were found for other education levels (no formal education, primary school, secondary school). |

**Table 3. Continued**
| Author (year) | Country | Sample | Exposure/outcome | Adjusted factors | Findings |
| --- | --- | --- | --- | --- | --- |
| Liang and Yu (2022) | China | 14,218 HH | EM: Age at first marriage (< 16, < 18 or <20/22 years<br>Educational attainment: Years of schooling completed | Parental years of schooling, parental age, gender, ethnic group, urban registration, migration status, parental absence in mid childhood, birth cohort, region, province. | A 1-year increase in schooling was associated with a lower likelihood of marriage before 16 and 18 years (coeff: -0.01 [SE: 0.01; p<0.01] and -0.02 [0.01; p<0.05] respectively) but not marriage before 20/22 years (0.02 [0.02]). |
| Marphatia et al. (2019) | India | 648 mother and child pairs | EM: Age at marriage < 18 years<br>Educational attainment: None to primary (1-5 years), upper primary or greater (≥6 years) | Sex, child growth, HH characteristics, maternal phenotype | EM associated with higher odds of secondary school dropout (OR: 1.50 [CI: 1.01, 2.25])<br>A greater proportion of girls dropped out from secondary school than boys and most likely due to getting married (70.8% vs 1.7%, p≤ 0.001) |
| Marphatia et al. (2020) | Nepal | 6,406 women aged 23-30 years | EM: Age at marriage ≤15 years (childhood), 16–17 years (adolescence) or ≥18 years<br>Educational attainment: none, 1–5 y (primary), 6–8 y (lower secondary), 9–10 y (secondary), or 11–13 y (higher secondary or above) | Husband's education, caste. | Women's later greater educational attainment was associated with age at marriage ≥18 years, though most strongly with 8 or more years of education (OR 1.03 [95% CI: 0.83,1.28], 1.27 [0.96-1.65], 2.22 [1.70-2.90], 5.02 [3.48-7.25] for 1-5 years, 6-8 years, 9-10 years and 11-13 years of education respectively vs none). |
| Paul (2019) | India | 122,955 women aged 20-24 years in 640 districts | EM: Age at first marriage<br>Educational attainment: No education/ illiterate, primary (1-5 years), secondary (5-10 years), higher secondary (10-12 years) college/ higher (12+ years) | Urbanization, religion (Hindu population), women autonomy (HH decision making, physical mobility, access to economic resources), and region. | Districts with a higher percentage of girls with no education/illiteracy and primary education had an increased risk of EM (coeff: 0.52 [SE: 0.03] and 0.66 [0.11] respectively with p < 0.01)<br>Districts with a higher percentage of girls with secondary, higher secondary, and college/higher education had a reduced risk of EM (-0.31 [0.06], -1.49 [0.09], and -1.31 [0.08] respectively with p <0.01 or 0.05) |
| Paul (2020) | India | 122,955 women aged 20-24 years | EM: Age at first marriage <18 years<br>Education level: Illiterate, primary, secondary, higher | Socio-economic characteristics of women (i.e., place of residence, caste, religion, education and wealth quintile) | Compared to illiterate women, the odds of EM decreased with an increase in education level:<br>primary school (OR: 0.87 [CI: 0.83, 0.92])<br>secondary school (0.53, [0.50, 0.55])<br>higher school (0.17 [0.16, 0.19]) |

**Table 3. Continued**
| Author (year) | Country | Sample | Exposure/outcome | Adjusted factors | Findings |
| --- | --- | --- | --- | --- | --- |
| Prakash et al. (2017) | India | 2,275 adolescent girls aged 13-14 | EM: Engaged/married<br>School dropout: currently in school (yes/no)<br>School absenteeism: Absent from school for 4+ days in past month were categorised as frequently absent | Cohort wave and district, age caste, female headed households, family wealth, family debt, family support for girls' education, number of siblings, older/younger sibling. | EM girls had higher odds of dropping out of school compared to never married girls (OR: 3.69 [CI: 2.12, 6.40]). |
| Roy and Chouhan (2021) | India | 357 women aged 15-49 years | EM: Age at marriage <18 years<br>Education level: Illiterate, primary, secondary, higher | Age, socioeconomic variables including religion, social group, parental education, father occupation, family income. | Compared to illiterate women, the odds of EM decreased with an increase in education level:<br>primary school (OR: 0.70 [CI: 0.32, 0.98])<br>secondary school (0.72 [0.36, 0.81])<br>higher school (0.06 [0.02, 0.19]) |
| Sagalova et al. (2021) | West and Central Africa (WCA) | 262,721 girls aged 15-19 years | EM: Age at marriage 10-14 years or 15-19 years<br>Educational attainment: None/ < primary, primary, incomplete secondary, secondary, or higher. | Cultural norms, average living standards | Across WCA, first marriage at age 10-14 years and ages 15-19 years associated with a higher likelihood of having educational attainment below primary school (coeff: 0.29 [SE: 0.003; p< 0.001] and 0.20 [0.002]; p<0.001). |
| Sekine & Hodgkin (2017) | Nepal | 1,631 girls aged 15-17 years | EM: Age at first marriage<br>School attendance: Yes, no<br>School dropout < secondary school (17 years): Yes, no | Age, urban/rural residence, religion, HH wealth, social class, HH head education. | Married girls had lower odds of school attendance (OR:0.10 [CI: 0.06, 0.17]; p<0.001) and dropout (10.04 [5.84, 17.25]; p<0.001) compared to those unmarried. |
| Singh, Shekhar & Gupta (2024) | India | 171,199 ever or currently married women aged 20–29 years | EM: Age at first marriage < 18 years<br>Educational status: No education, Primary, Secondary, and Higher. | Individual-level factors such as age, education, mass media exposure, and prior relationship with the husband, household-level factors (not specified) and household/community-level factors (not specified). | As education increases, the likelihood of early marriage decreases significantly. Women with primary education (OR: 0.93 [CI: 0.89, 0.97]), secondary education (0.53 [CI: 0.51, 0.54]), and higher education (0.12 [CI: 0.11, 0.12]) all have lower odds of early marriage compared to those with no formal education. |
| Zegeye et al. (2021) | Mali | 8,350 women aged 18-49 years | EM: Age at marriage < 18 years<br>Educational level: No formal education, primary school, secondary school, higher. | Husband education, women's and husband's occupation, economic status (ES), ethnicity, media exposure, family size, urban/rural residence, problematic distance to health facility, region, community literacy, community ES. | Having secondary and higher education associated with lower odds of EM (OR: 0.71 [CI: 0.60, 0.85] and 0.25 [0.14, 0.44] respectively; p<0.001) – but not having primary school education (1.02 [0.87, 1.20]), compared to having no formal education. |

Abbreviations: EM: Early Marriage, HH: Household, CI: Confidence Interval, PR: Prevalence Ratio, SD: Standard Deviation, OR: Odds Ratio, SE: Standard Error, Coeff: coefficient, HR: Hazard ratio, RRR: Relative Risk Ratio.

### H4: Preterm birth is associated with reduced cognitive function

All five systematic reviews found evidence of poorer cognitive function in children born preterm, compared to their term-born peers (**Table 4**). With the exception of one study from Belarus, included in the review of Chan et al. (Chan, Leong, Malouf, & Quigley, 2016) and two studies from India, included in the review of McBryde et al. (McBryde, Fitzallen, Liley, Taylor, & Bora, 2020), all the studies analysed were from high-income countries.

**Table 4.**
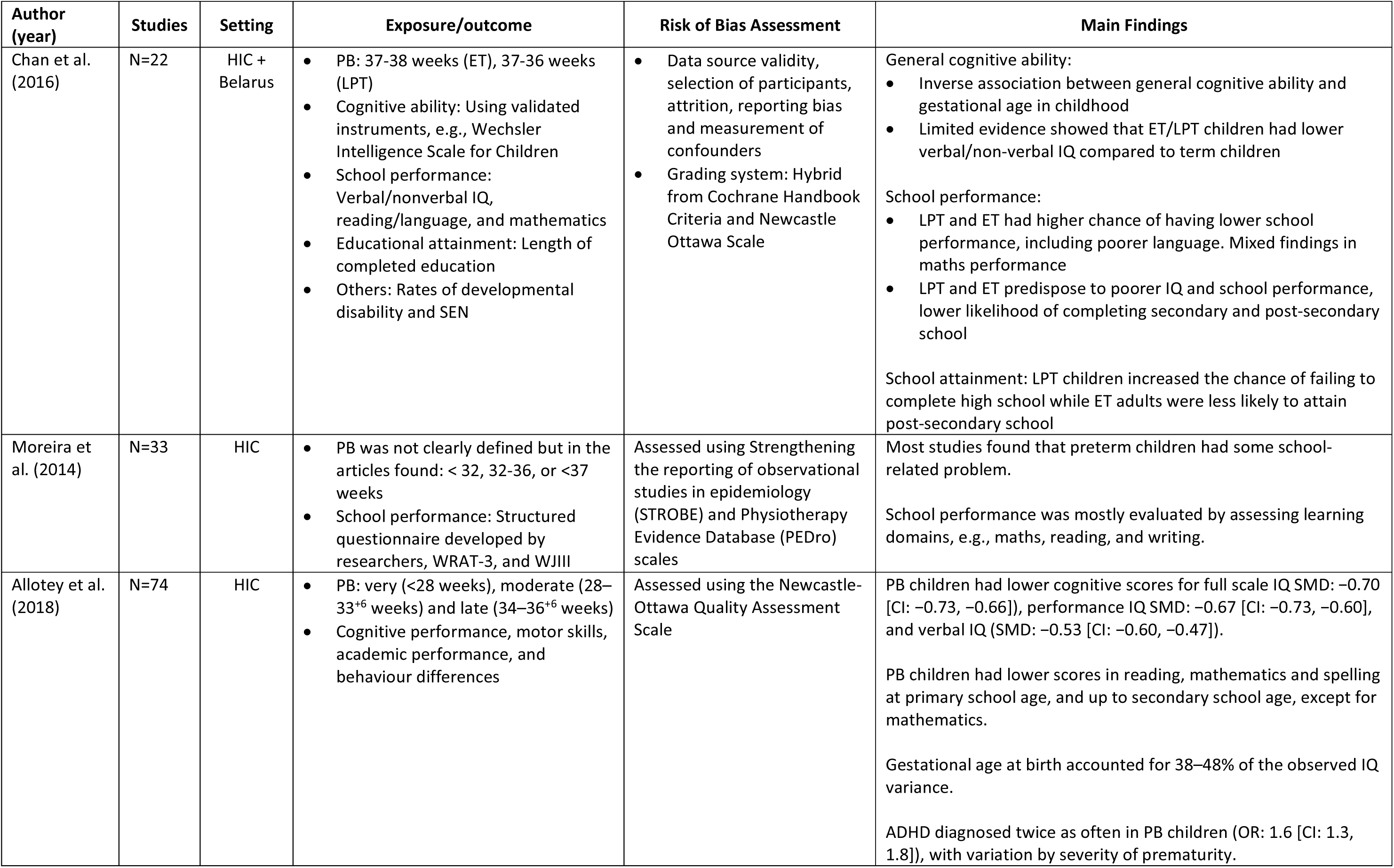
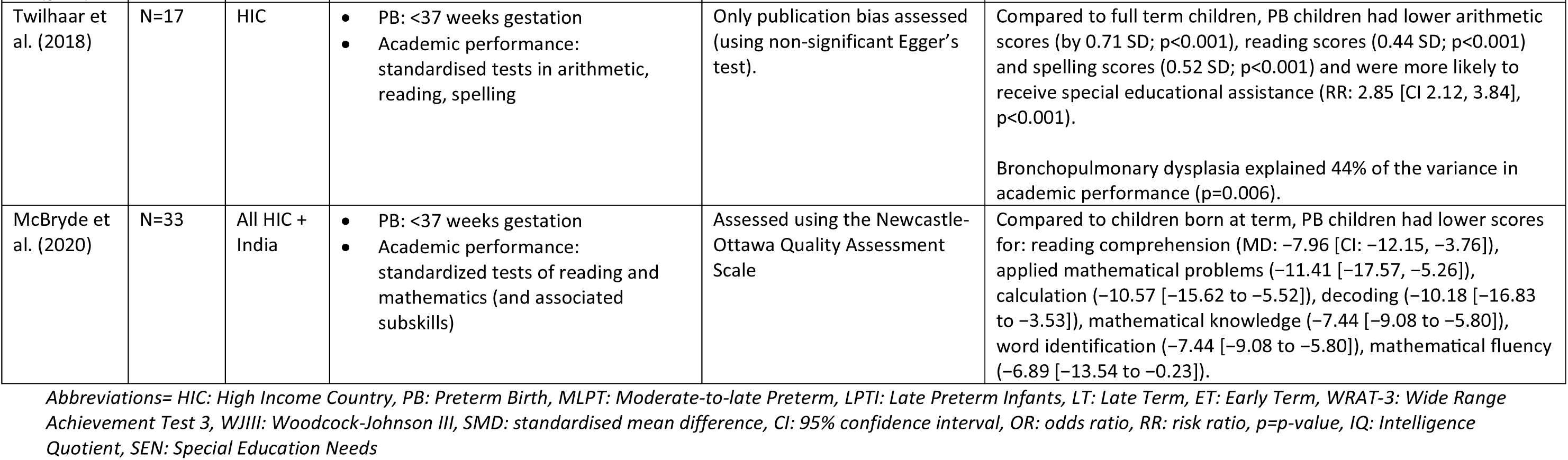
Summary of systematic reviews on the association of preterm birth with educational outcomes.

| Author (year) | Studies | Setting | Exposure/outcome | Risk of Bias Assessment | Main Findings |
| --- | --- | --- | --- | --- | --- |
| Chan et al. (2016) | N=22 | HIC + Belarus | • PB: 37-38 weeks (ET), 37-36 weeks (LPT)<br>• Cognitive ability: Using validated instruments, e.g., Wechsler Intelligence Scale for Children<br>• School performance: Verbal/nonverbal IQ, reading/language, and mathematics<br>• Educational attainment: Length of completed education<br>• Others: Rates of developmental disability and SEN | • Data source validity, selection of participants, attrition, reporting bias and measurement of confounders<br>• Grading system: Hybrid from Cochrane Handbook Criteria and Newcastle Ottawa Scale | General cognitive ability:<br>• Inverse association between general cognitive ability and gestational age in childhood<br>• Limited evidence showed that ET/LPT children had lower verbal/non-verbal IQ compared to term children<br>School performance:<br>• LPT and ET had higher chance of having lower school performance, including poorer language. Mixed findings in maths performance<br>• LPT and ET predispose to poorer IQ and school performance, lower likelihood of completing secondary and post-secondary school<br>School attainment: LPT children increased the chance of failing to complete high school while ET adults were less likely to attain post-secondary school |
| Moreira et al. (2014) | N=33 | HIC | • PB was not clearly defined but in the articles found: < 32, 32-36, or <37 weeks<br>• School performance: Structured questionnaire developed by researchers, WRAT-3, and WJIII | Assessed using Strengthening the reporting of observational studies in epidemiology (STROBE) and Physiotherapy Evidence Database (PEDro) scales | Most studies found that preterm children had some school-related problem.<br>School performance was mostly evaluated by assessing learning domains, e.g., maths, reading, and writing. |
| Allotey et al. (2018) | N=74 | HIC | • PB: very (<28 weeks), moderate (28–33<sup>+6</sup> weeks) and late (34–36<sup>+6</sup> weeks)<br>• Cognitive performance, motor skills, academic performance, and behaviour differences | Assessed using the Newcastle-Ottawa Quality Assessment Scale | PB children had lower cognitive scores for full scale IQ SMD: –0.70 [CI: –0.73, –0.66]), performance IQ SMD: –0.67 [CI: –0.73, –0.60], and verbal IQ (SMD: –0.53 [CI: –0.60, –0.47]).<br>PB children had lower scores in reading, mathematics and spelling at primary school age, and up to secondary school age, except for mathematics.<br>Gestational age at birth accounted for 38–48% of the observed IQ variance.<br>ADHD diagnosed twice as often in PB children (OR: 1.6 [CI: 1.3, 1.8]), with variation by severity of prematurity. |

**Table 4. Continued**
| Author (year) | Studies | Setting | Exposure/outcome | Risk of Bias Assessment | Main Findings |
| --- | --- | --- | --- | --- | --- |
| Twilhaar et al. (2018) | N=17 | HIC | • PB: <37 weeks gestation<br>• Academic performance: standardised tests in arithmetic, reading, spelling | Only publication bias assessed (using non-significant Egger's test). | Compared to full term children, PB children had lower arithmetic scores (by 0.71 SD; $p<0.001$), reading scores (0.44 SD; $p<0.001$) and spelling scores (0.52 SD; $p<0.001$) and were more likely to receive special educational assistance (RR: 2.85 [CI 2.12, 3.84], $p<0.001$).<br>Bronchopulmonary dysplasia explained 44% of the variance in academic performance ($p=0.006$). |
| McBryde et al. (2020) | N=33 | All HIC + India | • PB: <37 weeks gestation<br>• Academic performance: standardized tests of reading and mathematics (and associated subskills) | Assessed using the Newcastle-Ottawa Quality Assessment Scale | Compared to children born at term, PB children had lower scores for: reading comprehension (MD: -7.96 [CI: -12.15, -3.76]), applied mathematical problems (-11.41 [-17.57, -5.26]), calculation (-10.57 [-15.62 to -5.52]), decoding (-10.18 [-16.83 to -3.53]), mathematical knowledge (-7.44 [-9.08 to -5.80]), word identification (-7.44 [-9.08 to -5.80]), mathematical fluency (-6.89 [-13.54 to -0.23]). |

Abbreviations= HIC: High Income Country, PB: Preterm Birth, MLPT: Moderate-to-late Preterm, LPTI: Late Preterm Infants, LT: Late Term, ET: Early Term, WRAT-3: Wide Range Achievement Test 3, WJIII: Woodcock-Johnson III, SMD: standardised mean difference, CI: 95% confidence interval, OR: odds ratio, RR: risk ratio, $p$ = $p$ -value, IQ: Intelligence Quotient, SEN: Special Education Needs

Chan et al (Chan et al., 2016), analysing 22 studies, found evidence of an inverse association of gestational age with general cognitive ability, with limited evidence that late preterm and early term children had lower verbal and non-verbal IQ scores. Late preterm and early term children demonstrated poorer school performance, and reduced likelihood of completing secondary and post-secondary school. Moreira et al (Moreira, Magalhaes, & Alves, 2014), analysing 33 studies, found that preterm children had poorer academic performance in 15 of 16 studies. Allotey et al (Allotey et al., 2018), analysing 74 studies, found that children born preterm had lower IQ scores and lower scores in reading, mathematics and spelling at primary school age, and similar differences up to secondary school age, except for mathematics. Gestational age at birth accounted for 38–48% of the observed IQ variance. Twilhaar et al. (Twilhaar, de Kieviet, Aarnoudse-Moens, van Elburg, & Oosterlaan, 2018), analysing 17 studies, found that preterm children had lower ability in arithmetic, reading and spelling, and were 2.8 times (95% CI 2.1, 3.8) more likely to receive special educational assistance. McBryde et al. (McBryde et al., 2020), analysing 33 studies, found that children born preterm had lower scores for reading comprehension, word identification and mathematical abilities.

## Discussion

Studying intergenerational cycles of disadvantage is challenging, as the ideal approach would be a prospective longitudinal cohort that followed the second generation into adulthood and recorded their own reproductive outcomes. Such studies are rare, especially in low-income settings, hence we adopted an alternative approach, searching the literature systematically for evidence of the three steps that we hypothesised constitute an intergenerational cycle of risk linking early women’s marriage, preterm birth and school dropout.

Our review found that each part of the cycle has consistent supporting evidence, though relatively few studies to date have investigated whether early marriage is associated with the risk of preterm birth. Although studies from LMICs on the association of preterm birth with school dropout are also limited, there is substantial evidence from HICs that preterm birth undermines cognitive ability and schooling outcomes. Therefore, the concept of the intergenerational cycle appears valid. All three hypotheses have supporting evidence from India, though not within the same community.

An increased risk of preterm birth following early marriage was evident in four studies, two from South Asia where marriage is a near-universal practice, and two from South America where marriage is less universal. One underlying mechanism may involve early childbearing, as adolescent mothers have an increased risk of delivering a preterm offspring (Fall et al., 2015; Gronvik & Fossgard Sandoy, 2018). However, the study from Nepal is particularly informative, as it disentangled the risks of preterm birth associated with early marriage and early reproduction (Miller et al., 2022). The finding that the risk of preterm birth was elevated in primigravidae who had married very early (<14 years), independent of their age of childbearing, suggests that exposure to psychosocial stress might be part of the underlying mechanism. Further research is needed to understand how both early marriage and early reproduction relate to the risk of preterm birth.

The risk of school dropout in LMICs following preterm birth was demonstrated by two studies. A study of 700 adolescents from India showed that the risk of secondary school dropout decreased with each additional week of gestational age. A much larger study, combining data from five birth cohorts in Brazil, Guatemala, India, the Philippines and South Africa found that educational attainment was 0.44 years lower (95% CI 0.17, 0.71) following preterm birth compared to term birth. Systematic reviews of studies from any setting, the vast majority from high-income settings, supported the hypothesis that preterm birth impairs cognitive capacity and school performance, and increases the likelihood of school dropout. In addition, a large study from Scotland found a near-linear association of shorter gestation with difficulties in school, whereby each 1-week reduction in gestational length below 41 weeks was associated with an increased likelihood of the child requiring special educational need (MacKay et al., 2010). However, there are also plausible mechanisms that might contribute, for example, children born preterm may remain smaller than their age-peers, and might be held back from starting school, undermining their educational potential.

Finally, studies from across the Global South (mainly from sub-Saharan Africa and Asia) provided consistent evidence that school dropout is associated with increased risk of girls’ early marriage. While using heterogenous study designs, the studies were consistent in showing that school dropout was associated with increased risk of early marriage, and that early-married girls had lower educational attainment.

While the associations we identified are not determinative at the individual level, and refer rather to increased risks of adverse outcomes following the exposures, the population burden may nevertheless be substantial because each of preterm birth, school dropout and early marriage are common in the Global South. For example, a modelling study projected an assumed effect of preterm birth on educational attainment onto 622 million live births across five birth cohorts, spanning 121 countries (Blakstad et al., 2022). Across all countries combined, the model indicated that reducing preterm birth to a theoretical minimum prevalence of 5.5%, based on evidence from the INTERGROWTH-21^st^ study, would be associated with a potential gain of 9.8 million school years (95% CI: 1.5,18.4), including 3.66 (0.55,7.37) million years in South Asia and 3.06 (0.46, 5.92) million in Sub-Saharan Africa (Blakstad et al., 2022). These results are not expressed in a typical extended period in school at the individual level, but they indicate the potential for a change in one component of the cycle to impact another at scale.

The underlying mechanisms in the hypothesised cycle require further attention. As discussed above, early marriage may increase the risk of preterm birth through pathways such as psychosocial stress, inadequate nutrition, early reproduction or incomplete pelvic growth. The link between preterm birth and school dropout might involve direct detrimental effects on brain growth or function (MacKay et al., 2010), or it could reflect a common underlying driver. For example, a study by Huang et al. assessed heavy metal exposures in the cord blood of Bangladeshi babies and their associations with preterm birth. Titanium, arsenic and barium exposure all predicted preterm birth, with an increased element risk score almost tripling the odds of preterm birth (OR=2.72, 95% CI: 1.57-4.69) (Huang et al., 2021). These metals may also impair brain development (C. Liang et al., 2020). Interestingly, Huang et al. found a significant moderation effect of child marriage on their element risk score; an association between cord blood element load and preterm birth was only found in women who married before 18 years. A study by Rahman et al. also conducted in Bangladesh found that both early marriage and arsenic exposure were associated with preterm birth, and that a lowering of pregnancy weight gain mediated these associations (Rahman et al., 2018). Therefore, early marriage may increase susceptibility to other biological stressors.

Finally, although school dropout may precipitate early marriage (as families may elect to marry a daughter early following poor school performance), the reverse scenario may also occur, as girls may be prevented from attending school following their early marriage. A study from India found evidence for both pathways, and also that a small minority of early married girls were still attending school (Marphatia et al., 2022). Moreover, schooling in some societies is directly related to marriage decisions, as greater education increases the value of an incoming bride to the marital household, and also affects the amount of dowry (Jeffery & Jeffery, 1994). Therefore, the association of educational attainment and marriage is complex and merits further attention.

Importantly, common factors precipitating both school dropout and early marriage may also lie outside the family and household domain. For example, in the Democratic Republic of Congo, civil conflict drove girls to drop out of school, which was in turn associated with earlier sexual debut and adolescent marriages, and ultimately adolescent motherhood (Mugisho, 2024).

Our findings are especially relevant to settings such as South Asia where marriage is near-universal, as efforts to delay marriage have the greatest potential to disrupt this intergenerational cycle. In South America, where marriage is less obligatory, unmarried women have an increased risk of having a child born preterm compared to married women (M. L. Urquia et al., 2022; Marcelo L. Urquia et al., 2022), whereas among married women, earlier marriage was associated with increased risk.

Our findings are consistent with the hypothesis that early marriage depletes maternal capital, an umbrella term for components of maternal phenotype that promote the capacity for investment in offspring (J.C. Wells, 2010). While the evidence that we have reviewed supports the intergenerational cycle that we hypothesised, in reality the components we focused on are part of a broader intergenerational cycle that also includes the detrimental effects of malnutrition and poverty on maternal capital. In Brazil and India, we have shown that depletions in both biological and social components of maternal capital impair outcomes of the offspring, and moreover increase the likelihood of the same depletions in maternal capital recurring in the next generation, when the offspring reach adulthood and start reproducing (Marphatia, Wells, Reid, Bhalerao, et al., 2024; J.C. Wells et al., 2019). Complementary to the pathways we have explored here, early marriage has also been associated with infant undernutrition (Raj et al., 2010; J.C. Wells et al., 2022), which in turn has been associated with school dropout (Katoch, Shrikant, Nawaz, Parihar, & Ahmed, 2022).

Our review adds to growing awareness of the intricate links between behaviours that might appear strongly cultural (household decisions about schooling and marriage), and biological traits (brain development) that are sensitive to diverse physical factors such as malnutrition, pollution and the stress response. We need to move beyond disciplinary silos, whereby education is seen only as a school-based issue, and marriage as only a household transactional issue, to understand that the variability in such decisions has much deeper roots that are embedded in biological mechanisms.

Our study had some strengths, including the use of systematic searches to obtain all relevant evidence, and the use of specific search terms that were able to identify a number of studies in both biomedical and social science literature. However, our approach also had some limitations. The number of available publications was low for H1 and H2, and we cannot demonstrate causality in the associations, though the consistency of findings across settings suggests that there may be a causal association.

## Conclusion

We found evidence from LMICs for each of the three steps in a biosocial intergenerational cycle of risk – that girls’ early marriage may increase risk of preterm birth, that preterm birth may increase risk of school dropout, in part through effects on cognitive function, and that school dropout may increase risk of girls’ early marriage. We propose that these relationships constitute a specific component of a broader inter-generational cycle, involving a larger number of traits and mechanisms

Breaking such intergenerational cycles will require political will, as the full benefits will inevitably take time to emerge. For this particular pathway, as preterm birth is difficult to prevent at the individual level, the more promising opportunities lie in promoting girls’ education across the whole range of educational ability, and preventing early marriage.

## Data Availability

Data sharing not applicable to this article as no datasets were generated or analysed during the current study.

## Ethics statement

Not applicable

